# radioGWAS: link radiome to genome to discover driver genes with somatic mutations for heterogeneous tumor image phenotype in pancreatic cancer

**DOI:** 10.1101/2023.11.02.23297995

**Authors:** Dandan Zheng, Paul M. Grandgenett, Qi Zhang, Michael Baine, Yu Shi, Qian Du, Xiaoying Liang, Jeffrey Wong, Subhan Iqbal, Kiersten Preuss, Ahsan Kamal, Hongfeng Yu, Huijing Du, Michael A. Hollingsworth, Chi Zhang

## Abstract

Addressing the significant level of variability exhibited by pancreatic cancer necessitates the adoption of a systems biology approach that integrates molecular data, biological properties of the tumors, and clinical features of the patients. In this study, a comprehensive multi-omics methodology was employed to examine a distinctive collection patient dataset containing rapid autopsy tumor and normal tissue samples as well as longitudinal imaging with a focus on pancreatic cancer. By performing a whole exome sequencing analysis on tumor and normal tissues to identify somatic gene variants and a radiomics feature analysis to tumor CT images, the genome-wide association approach established a connection between pancreatic cancer driver genes and relevant radiomics features, enabling a thorough and quantitative assessment of the heterogeneity of pancreatic tumors. The significant association between sets of genes and radiomics features revealed the involvement of genes in shaping tumor morphological heterogeneity. Some results of the association established a connection between the molecular level mechanism and their outcomes at the level of tumor structural heterogeneity. Because tumor structure and tumor structural heterogeneity are related to the patients’ overall survival, patients who had pancreatic cancer driver gene mutations with an association to a certain radiomics feature have been observed to experience worse survival rates than cases without these somatic mutations. Furthermore, the outcome of the association analysis has revealed potential gene mutations and radiomics feature candidates that warrant further investigation in future research endeavors.

## Introduction

Pancreatic ductal adenocarcinoma (PDAC) is a critical global health problem, with a mortality rate remaining the highest among major solid cancers. Despite decades of clinical and research efforts, the 1-year survival rate is 20% and the 5-year survival rate remained single digit for many years and only recently rose to 10%^1,2^. Clearly, new and synergistic approaches are needed to battle this ferocious disease.

Pancreatic cancer is often detected at late stages and has a weak response to current chemotherapy and a poor overall prognosis. Some hereditary risks were discovered, suggesting that up to 15% of pancreas cancer is attributable to genetic causes^3^. Genomic analysis plays an important role in understanding the complex biology of pancreatic cancer development and progression, and in identifying novel treatments targeting specific molecular pathways. However, the hallmark of pancreatic cancer is a high degree of heterogeneity in the biology of pancreatic tumor progression. Clonal variations were observed in premalignant and malignant tumors that result in different and multiple biological properties of tumors that progress to kill the patient^4,5^. These differences are manifested as tumors that progress with different biological properties, which affects the nature of the cells that grow and metastasize and the capacity of these cells to influence and organize their tumor microenvironment. This heterogeneity can be seen both at the molecular level, with non-consensus mutations and gene expression patterns, at the histological level, with different cell types and structures within the tumor, or at the tumor imaging level, with various appearances on CT images^6,7^. For the molecular level of intertumoral heterogeneity, whole exome sequencing analyses revealed a complex mutational landscape for PDAC^8^. Although mutations of some genes, such as KRAS and TP53, occur at rates of up to >50%, the frequency of most other recurrently mutated genes is less than 10% and there is a long tail of infrequently mutated genes among PDAC patient population^9,10^. This degree of heterogeneity has previously been underestimated or understated in the literature and in studies that undertake the discovery of biomarkers related to disease progression. It is also reflected by the results of population-based DNA sequencing and RNA expression studies to date, in that no consistent pattern of mutations or RNA expression profiles have yet been defined that accurately predict biological aspects of disease progression^11,12^.

Tackling this high degree of heterogeneity of pancreatic cancer demands a system science approach that integrates molecular data, biological properties of the tumors, and clinical features of the patients. The quantitative approaches for medical imaging analysis, such as extracting radiomics features, are perfect for globally assessing the heterogeneity of PDAC at the tumor imaging level^13^. In this work, we present a radiome-wide and genome-wide association approach to identify the driver genes for the heterogenicity at the tumor phenotype level. This method was conducted in a unique patient population for which a large amount of tumor and normal tissue samples were collected in a rapid autopsy immediately following patient’s demise. Our work demonstrates the feasibility of this novel systematic approach to provide new insight into the molecular mechanisms of pancreatic cancer progression.

Currently, whole exome sequencing (WES) is widely used to identify cancer-driver genes by searching for genes with a high rate of somatic mutation recurrence in multiple patient samples. What roles do these WES-identified cancer driver genes play in intertumor heterogeneity? This is the scientific question that we wish to answer. We collected a cohort of PDAC patients, who had both tumor and healthy tissues from rapid autopsy, and pancreatic contrast-enhanced CT images. WES was conducted on tumor tissues and healthy tissues. After conducting a comprehensive WES analysis and a tumor image radiomics analysis on these patients, we performed an association study to identify the cancer-driver genes that are significantly associated with image features. Our results shed some new light on tumor genomic and morphological heterogeneity for PDAC.

## Results

### Patient and dataset information

The patient and tumor clinical characteristics of our studied cohort are listed in Table 1. The patients in the cohort had a median of 6 (range: 3-30) serial pancreas contrast CTs available. The CT used for analysis, i.e. the last CT in the series for each patient was acquired a median of 34 (range: 1-579) days before the date of death.

**Table 1.** Patient and tumor clinical characteristics.

| Characteristic | Number of Patients (Percentage)<br>Median (Range) |
| --- | --- |
| **Gender** |  |
| Male | 16 (60%) |
| Female | 10 (40%) |
| **Median age (range)** | 71 (40-89) |
| **Tumor site in pancreas** |  |
| Head | 20 (77%) |
| Neck | 1 (4%) |
| Tail | 1 (4%) |
| Body | 4 (15%) |
| **Stage at diagnosis** |  |
| II | 9 (35%) |
| III | 4 (15%) |
| IV | 13 (50%) |
| **Use of radiotherapy** |  |
| Yes | 8 (31%) |
| No | 18 (69%) |
| **Use of chemotherapy** |  |
| Yes | 25 (96%) |
| No | 1 (4%) |
| **Survival days** | 331 (27-2282) |

### Somatic single-nucleotide variants (SNVs)

Driver somatic SNVs are genetic changes in a cell that drive the development and progression of cancer. In pancreatic cancer, several driver mutations have been identified that contribute to the development of the disease ^14,15^. In this study, we only retained translationally consequential SNVs, i.e. the missense variant, stop codon gain, start codon lost, sequence feature, splice donor variant, and intron variant. Detailed information on these somatic SNVs obtained from WES for this population is shown in Supplementary Table S1. The number of SNV recurrences in the above categories was counted for the patient population. Based on the single SNV recurrence, the mutations Chr12:25245350 C->T|A (G12D and G12V mutations) on KRAS had the highest recurrence rate, in 14 out of 26 patients (54%). These mutations, G12D and G12V, have been reported as the most common ones in pancreatic cancer, recurring at about 45% and 35%, respectively ^16,17^. The second largest recurrence is 9 (35%), for the SNV Chr12:25245351 C->G|A (G12R and G12C mutations) also on KRAS. These two SNVs caused the same missense variant on the amino acid sequence. Subsequently, the SNVs, Chr2:130074357 on *POTEF* and Chr7:152358679 on *KMT2C* had 7 (27%) and 6 (23%) recurrences out of 26 patients, respectively. For each gene, we also calculated the number of individuals carrying variants in any given gene. The top-ranked genes are *KRAS* (22), *TP53* (17), *KMT2C* (17), *LRP1B* (14), *FGFR2* (13), *RGPD3* (11), *EWSR1* (10), and *RGPD4* (10) (Supplementary Table S2).

The *KRAS* gene had SNVs in 22 out of all 26 patients (84.6%). This agrees with the discovery that KRAS mutations are found in more than 90% of PDACs^18-20^. For gene *TP53*, 17 out of 26 patients (65.4%) had mutations, and previous works showed that *TP53* mutations were found in approximately 50% of PDACs and are associated with a poor prognosis. 17 out of 26 patients (65.4%) also had *KMT2C* mutations. Histone Lysine Methyltransferase (KMT2) family genes are frequently mutated in multiple cancer types ^21^. Histone Lysine Methyltransferase 2C (KMT2C), also known as myeloid/lymphoid or mixed-lineage leukemia protein 3 (MLL3) is among the most frequently mutated cancer genes in major cancer types^22,23^. These genes have somatic mutations in most PDACs.

Gene *CDKN2A* had SNVs in 9 out of all 26 patients (34.6%). Previous studies showed that CDKN2A mutations are found in approximately 29% of PDACs and are associated with a poorer prognosis^24^. CDKN2A is a tumor suppressor gene that encodes the p16INK4A protein (hereafter mentioned as CDKN2A). As in its name, CDKN2A is a negative regulator of cell cycle progression (G1-to-S phase transition) by disturbing the complex formation between CDK4/6 and cyclin D^25,26^.

### Radiomics features

Radiomics features can be used to capture the heterogeneity of the tumor phenotype in the context of pancreatic cancer. Texture analysis can be used to quantify the spatial arrangement of pixel intensities within the tumor. With the close-to-a-thousand radiomics features extracted from each tumor VOI, a heatmap was generated to show the tumor radiomics pattern of the studied population (Figure 1). The detailed data of radiomics features for patients in this cohort are listed in Supplementary Table S3. The radiomics pattern did not show direct correlations with patient clinical data listed in Table 1. After feature selection, 170 radiomics features were kept. Out of the 170 radiomics features, 59 features had a coefficient of variation (CV), the ratio of the standard deviation to the mean, greater than 1, indicating higher heterogeneity among the studied population. Radiomics feature wavelet.LHH.firstorder.Skewness had the largest CV, at 35.9. Figure 2 shows 3 tumor images with different values of wavelet.LHH.firstorder.Skewness. This feature is related to the asymmetry of the distribution of pixel values after applying a wavelet filter. The large CV indicates the high heterogeneity of tumor pixel gray level distribution. The smallest CV including glcm.Idmn (5.98×10^−3^), glcm.InverseVariance (4.04×10^−2^), and original.shape.Sphericity (9.65×10^−2^), indicating high homogeneity of these features among all patients. Feature IDMN (inverse difference moment normalized) is used to assess the local homogeneity of VOI. Because all tumor images have very low local homogeneity, the values of IDMN of all tumor images were high, >0.98. Sphericity is a measurement of the roundness of the tumor region’s morphology relative to a sphere. It is a measure without dimensions, independent of scale and orientation. The majority of tumors in this data set are in their later stages, and hence, the tumors are not in a round shape.

**Figure 1.**
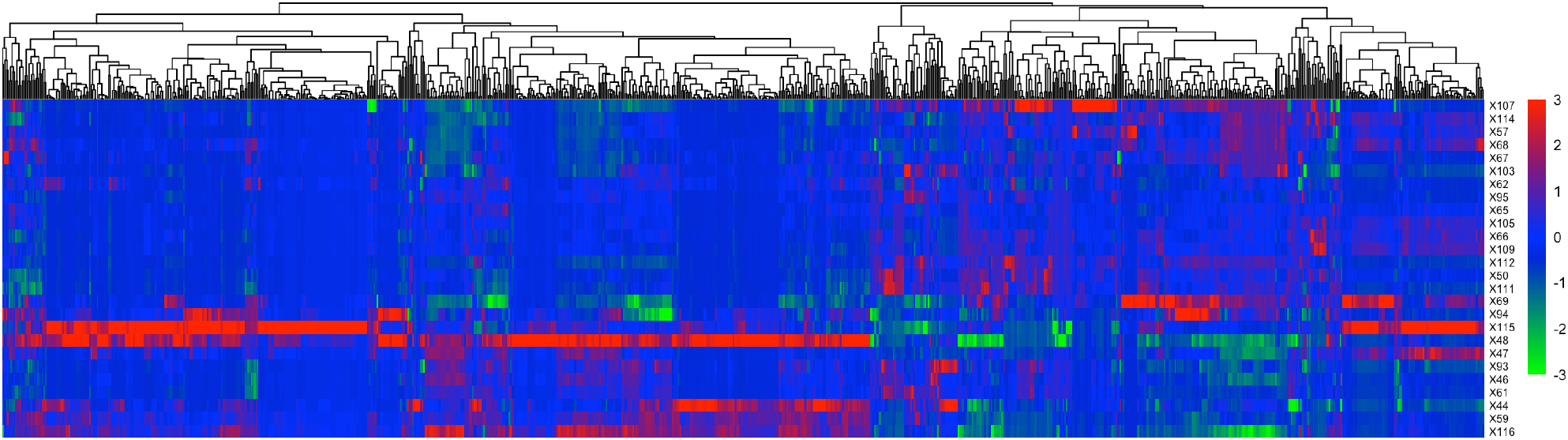
Radiomics features. Each tumor from a patient (row) has 944 radomics features (column). The color indicates the Z-score of a patient for a given radiomics feature.

**Figure 2.**
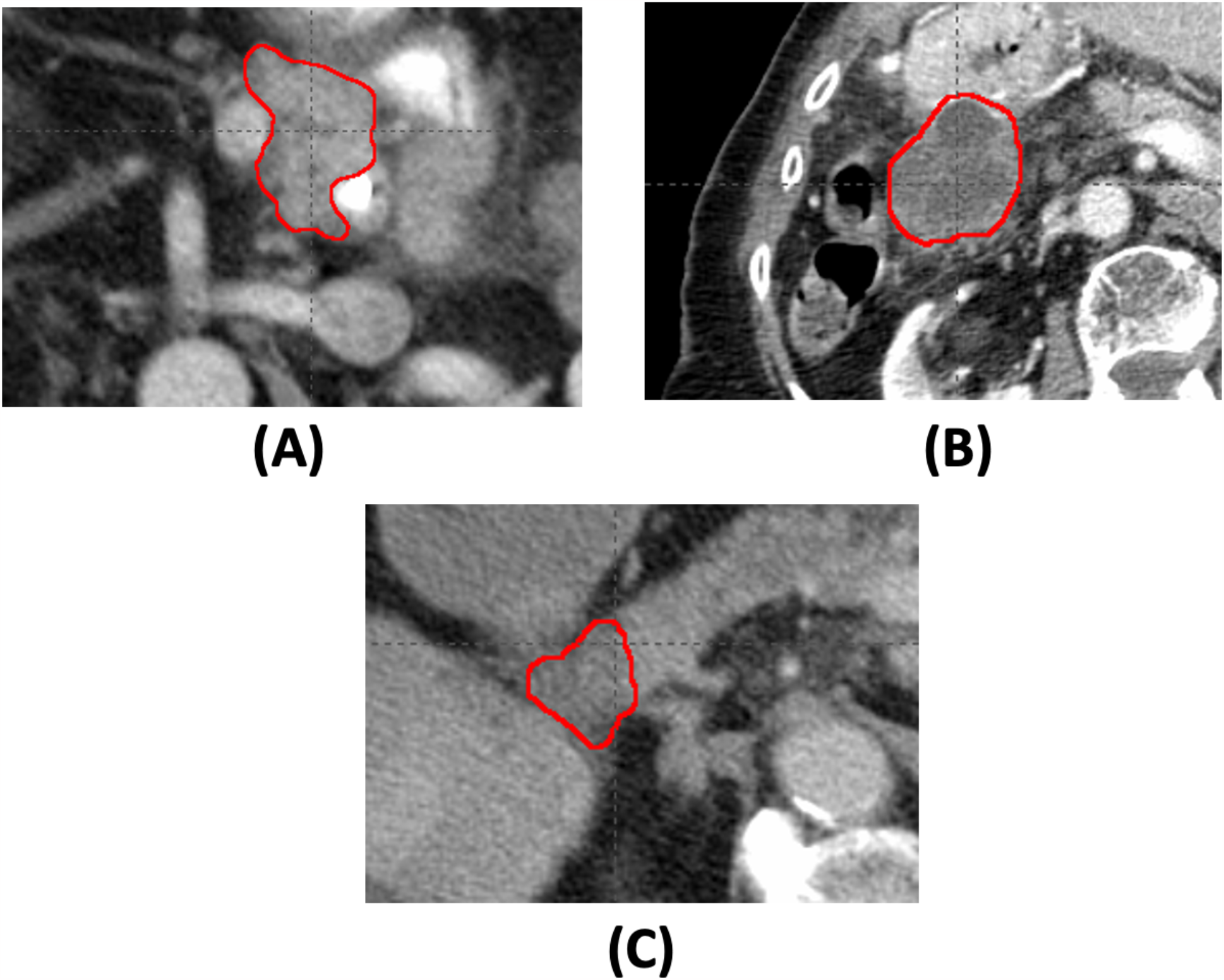
Example tumor images of three patients. The corresponding tumor images have the value of the radiomics feature, wavelet.LHH.firstorder.Skewness, −4.14 (A), −2.31×10^−3^ (B), and 1.31 (C), respectively.

### Radiome-wide and genome-wide association

This association analysis found several significant associations (P-value < 10^−4^) between radiomics features and genes with many somatic variants in the tumors. Table 2 lists the radiomics features and somatic variants with significant associations. Interestingly, there was no significant association found between radiomics features and driver genes with high recurrence frequency of somatic variants, such as *KRAS* and *TP53*. Genes that had a significant association with radiomics features have a recurrent rate of 3 (11.5%) to 7 (26.9%) among the patients.

**Table 2.**
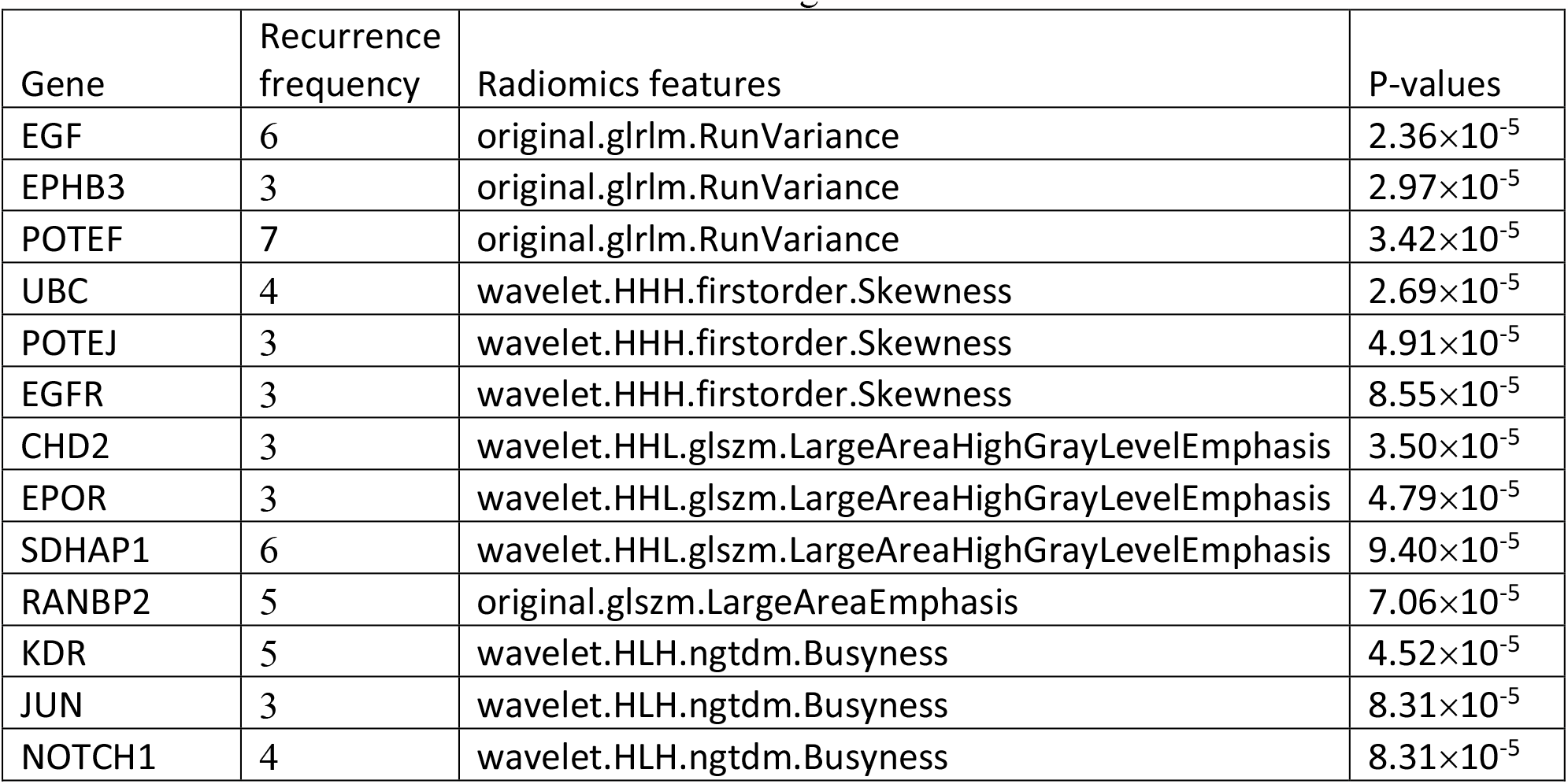
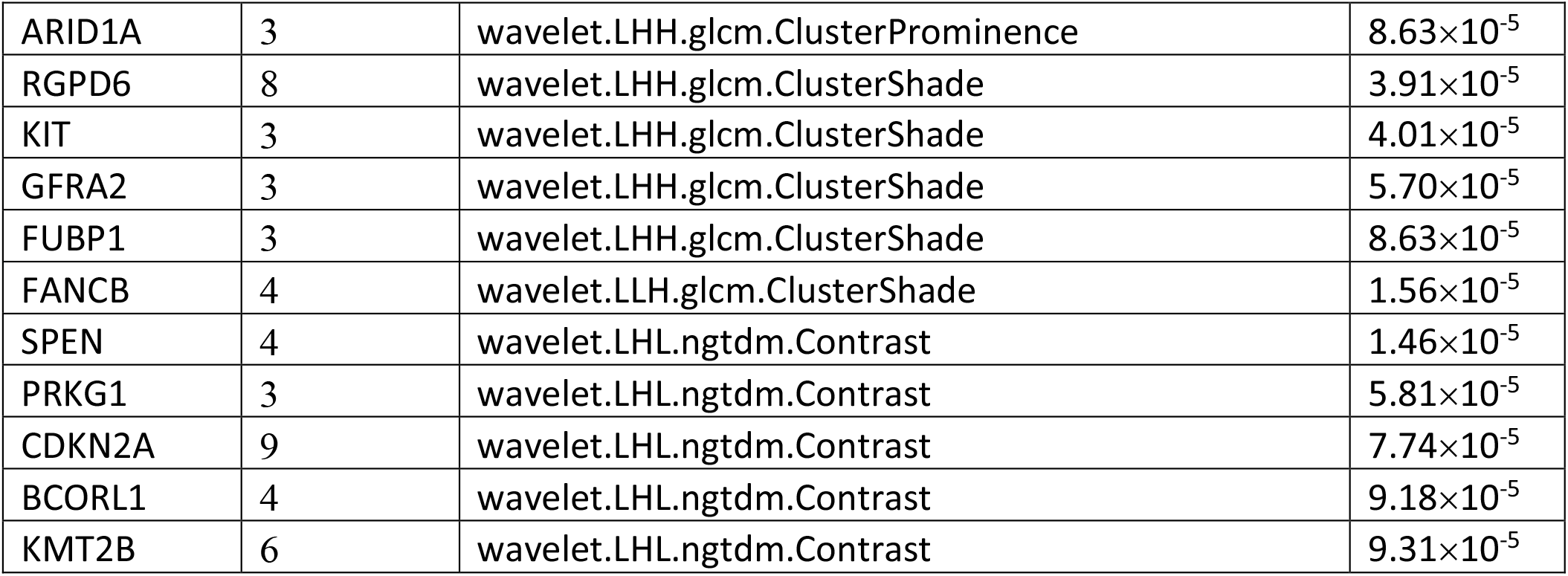
Associations between genes and radiomics features.

These genes that are significantly associated with the radiomics features are related to tumor formation and progress. For example, *EGF* and *EGFR* genes are important for cancer cell proliferation and spread in the body ^27,28^. The *EGFR* gene with somatic SNVs is associated with wavelet.HHH.firstorder.Skewness which indicates the tumor pixel value distribution asymmetry on the CT images. Mutations in the *EGFR* gene, which encodes epidermal growth factor receptors that enables cancer cells to grow and proliferate. The expression and function of the mutant *EGFR* gene may contribute to the varying patterns of tumor growth leading to the distinct skewness among patients. Some transcription factor genes, like RGPD6 which was the most commonly mutated in other types of cancers^29^, also had a significant association with the radiomics feature, wavelet.LHH.glcm.ClusterShade, which is a descriptor of tumor texture pattern.

Several genes are associated with the same radiomics feature. This indicates these genes may be involved in the same biological pathway or are involved in synergistic interactions. For example, *NOTCH-1, JUN*, and *KDR* genes were all associated with the radiomics feature, wavelet.HLH.ngtdm.Busyness. Literature suggests that both *Notch* and *JUN* genes are related to the cell apoptosis^30^, and Notch-1 promotes JNK/c-Jun activation^31^. Loss of function of either JUN or NOTCH-1 can result in similar observed biological effects. In our cohort, only one patient carried somatic mutations in both the *JUN* and *NOTCH-1* genes, suggesting that the two mutations may be alternative. Gene *KDR* encodes the Kinase insert domain receptor, also known as vascular endothelial growth factor receptor 2 (VEGFR-2). The corresponding radiomics feature, ngtdm, is a Neighboring Gray Tone Difference Matrix that quantifies the difference between a gray value and the average gray value of its neighbor pixels. Busyness is a measure of the change from a pixel to its neighbor. A high value for busyness indicates rapid changes in intensity between pixels and its neighborhood. This indicates that somatic SNVs in *NOTCH-1, JUN*, and *KDR* genes may cause cancer cells to develop at various speeds and result in varying localized colonization of different cell types in the tumor.

Three genes, *EGF, EPHB3*, and *POTEF*, were all associated with original.glrlm.RunVariance. Three patients have SNVs in at least two of these three genes. This is also a texture feature. A gray level run length is defined as the number of consecutive pixels that have the same gray level value. The gray level run length matrix (GLRM) consists of all run lengths in a VOI. Run Variance is the measure of the variance in runs for the run lengths, and hence, is related to the intratumor heterogeneity. As an example, Figure 3 shows the images of two tumors with or without an *EGF* gene mutation. The association between these three genes, *EGF, EPHB3*, and *POTEF*, and Run Variance indicates their roles in the heterogeneous colonization of cancer cells. For example, the gene, *EPHB3*, has been found to be involved in the signaling conduction for colonizing cells^32^.

**Figure 3.**
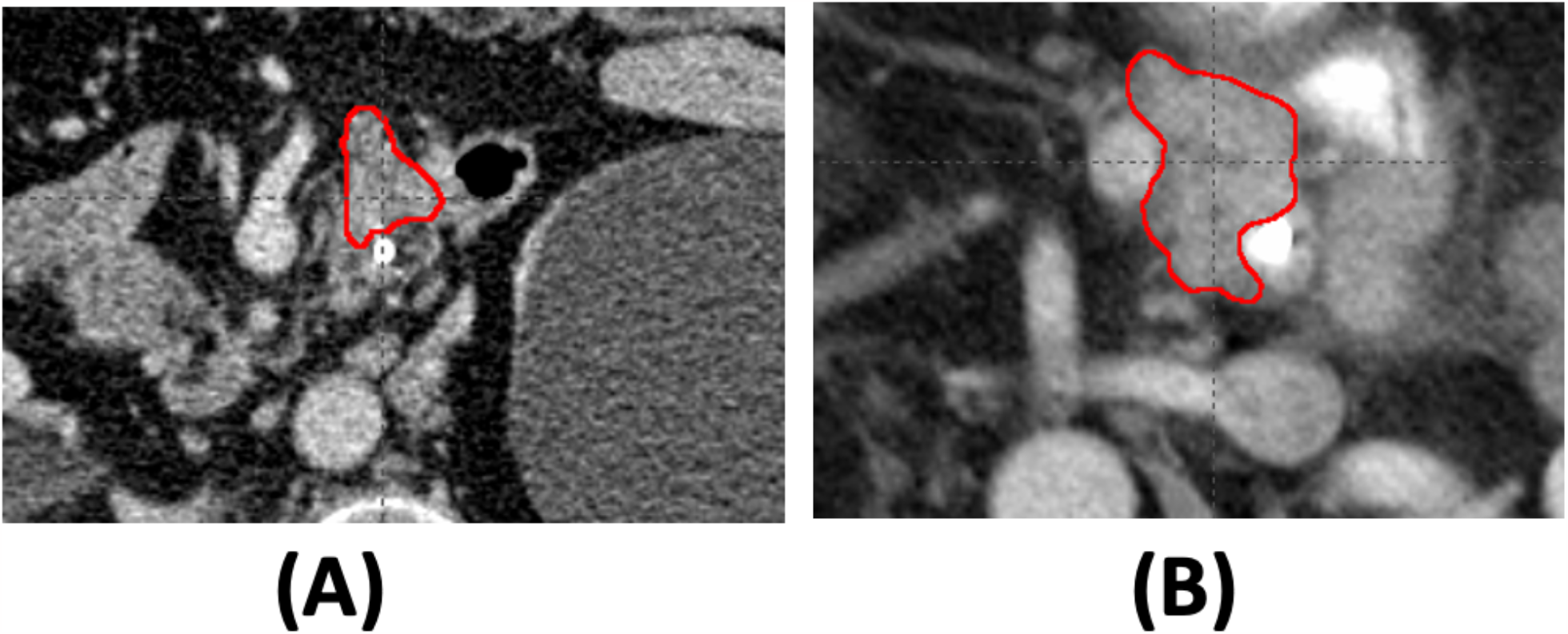
Tumor image of two different patients with (A) and without (B) a *SPEN* gene mutation. The CT scans were obtained from two patients, one of whom had SNVs on the *SPEN* gene, whereas the other did not. The corresponding tumor images have the value of the radiomics feature, wavelet.LHL.ngtdm.Contrast, 0.63 (A) and 3.02×10^−2^ (B), respectively.

Genes, including *SPEN, PRKG1, CDKN2A, BCORL1*, and *KMT2B*, had a significant association with wavelet.LHL.ngtdm.Contrast. This is also a texture feature. The Neighboring Gray Tone Difference (NGTD) is the difference between a gray value and the average gray value of its neighbors within a distance. The value of Contrast is a measure of the local intensity variation and the spatial intensity change. Figure 4 shows an example of the tumor CT images of two patients with or without the SNV in the *SPEN* gene. Its high value indicates that the tumor region has large changes between voxels and their neighborhood. This association suggests that these five genes are involved in tumor growth and tumor shape regulation. Especially, the gene *CDKN2A* has somatic SNVs in 9 patients (34.6%) out of 26 patients. The gene *CDKN2A*, whose gene product is the cyclin-dependent kinase inhibitor 2A, plays an important role in cell cycle regulation and demonstrates tumor suppressor activity. Inactivation of CDKN2A leads to uncontrolled cell growth^33^.

**Figure 4.**
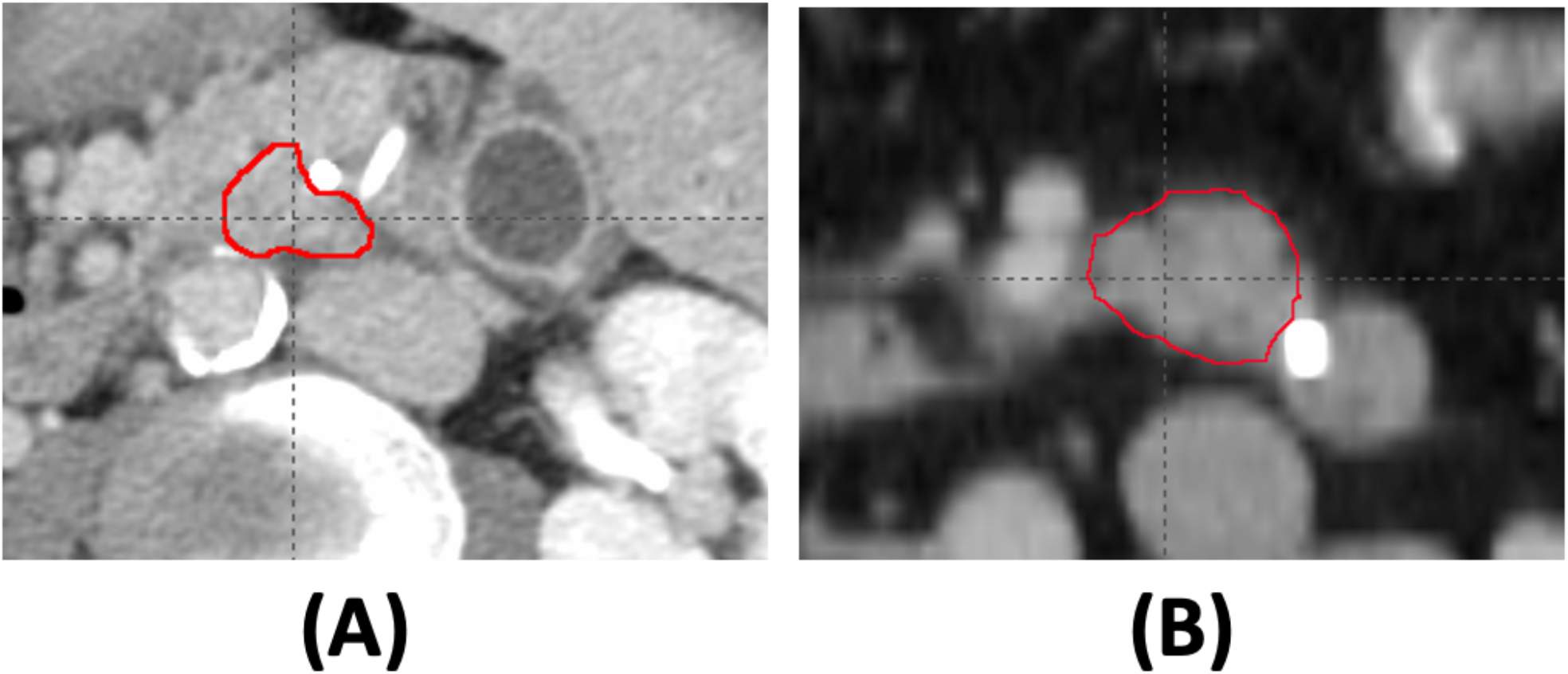
Tumor image of two different patients with (A) and without (B) an *EGF* gene mutation. The CT scans were obtained from two patients, one of whom had SNVs on the *EGF* gene, whereas the other did not. The corresponding tumor images have the value of the radiomics feature, original.glrlm.RunVariance, 0.26 (A) and 0.99 (B), respectively.

## Discussion

Previous whole-genome sequencing and variation analysis discovered that mutations on genes, *KRAS, TP53, CDKN2A, ARID1A, ROBO2, KDM6A*, and *PREX2*, are important in the pancreatic cancer^34^. Activating mutations of KRAS are nearly ubiquitous, being found in more than 90% of PDACs^18-20^. Among all cancers, KRAS mutations are present in ∼25% of tumors^35^and frequently in lung, colorectal, and pancreatic cancers^18,36-38^. Actually, the RAS family, including KRAS, NRAS, and HRAS, is the most frequently mutated gene family in all different types of cancers ^16^. The inactivation of TP53 reoccurs at rates of > 50% in pancreatic cancer. The gene, *TP53*, is the most frequently mutated gene in different cancers at rates from 38%–50%, such as ovarian, esophageal, colorectal, head and neck, larynx, lung, and pancreatic cancers ^39^. These genes, *KRAS* and *TP53*, are important in the formation of cancer and hence have been extensively studied before, but they are not often directly related to intertumor or intratumor heterogeneity. In pancreatic cancer, the prevalence of recurrently mutated genes then drops to ∼10% for a handful of genes involved in chromatin modification, DNA damage repair, and other mechanisms resulting in significant intertumoral heterogeneity ^34^. These driver mutations can be used to inform diagnostic and treatment strategies for pancreatic cancer and provide a better understanding of the underlying biology of the disease. To understand the role of these genes with low recurrence mutations, the association between gene-mutation profiles and radiomics features was conducted in this work. The significant association between sets of genes and radiomics features identified genes that contribute to morphological heterogeneity. These genes include *NOTCH-1, JUN, KDR, EGF, EPHB3, POTEF, SPEN, PRKG1, CDKN2A, BCORL1*, and *KMT2B*.

This work discovered that several genes with mutations have significant associations with the same radiomics features. Some genes associated with the same radiomics feature are in the same regulation pathway and work together for a regulatory cascade. For example, *Notch* and *JUN* are associated with wavelet.HLH.ngtdm.Busyness and the gene product Notch-1 promotes JNK/c-Jun activation^30,31^. Loss of function of either JUN or NOTCH-1 can result in cells evading cellular death pathways. Gene *JUN* had somatic mutations in three patients, and *NOTCH-1* gene had somatic mutations in four patients. Only one patient had mutations in both *JUN* or *NOTCH-1* genes, and this patient had 220 somatic mutations, which is much more than the average number (51) of somatic mutations per patient in our dataset. In contrast to these alternative genes, the other genes that were associated with the same radiomics feature appear to work in different pathways, and several somatic mutations in multiple genes need to work together. For example, EGF, EPHB3, and POTEF all are associated with original.glrlm.RunVariance, but EGF works for cell proliferation and EPHB3 works for colonizing cells. In this case, SNVs in multiple genes have a high chance to occur in the same patient. The knowledge of various recurrence patterns of cancer driver genes associated with a specific phenotypic feature has the potential to guide the combination therapy for cancer using multi-target medicines, which has recently garnered a great deal of attention as one of the most promising cancer-fighting tools^40,41^. And in our novel approach, radiomics features expand the big data space that we could integrate and leverage for novel discoveries.

Tumor heterogeneity can refer to intratumor heterogeneity, including heterogeneity of structures within a single tumor, or intertumor heterogeneity if tumors are compared among patients. The tumor structural heterogeneity can be quantified by medical images^42^, especially via radiomics features^43,44^. Radiomics features, such as tumor shape and texture features can be used to quantify tumor heterogeneity, which makes them the potential to serve as imaging-based heterogeneity biomarkers^45^. Texture analysis can be used to quantify the spatial arrangement of pixel intensities within the tumor. Usually, tumor image textural features could be extracted in several different ways, such as using GLRM-based approaches. In this study, we found a significant association between pancreatic cancer driver genes, *EGF, EPHB3*, and *POTEF*, and original.glrlm.RunVariance. This association links the molecular level mechanism and their outcomes at the level of tumor structural heterogeneity. Because tumor structure and tumor structural heterogeneity are related to the patients’ overall survival, patients who had pancreatic cancer driver gene mutations with an association to a certain radiomics feature showed worse survival rates than cases without those somatic mutations. For example, we used a public cohort of pancreatic cancer in the TCGA database^46^ and collected the survival information of patients with somatic mutation on genes, *CDKN2A, PRKG1*, and *BCORL1*, which had a significant association with wavelet.LHL.ngtdm.Contrast from our study. Figure 5 shows the survival curves comparing the patients with somatic mutation on genes, *CDKN2A, PRKG1*, and *BCORL1*, and the other patients without. The survival time of patients with somatic mutation on these driver genes is shorter than the other patients (FDR-adjusted P-value = 0.0348).

**Figure 5.**
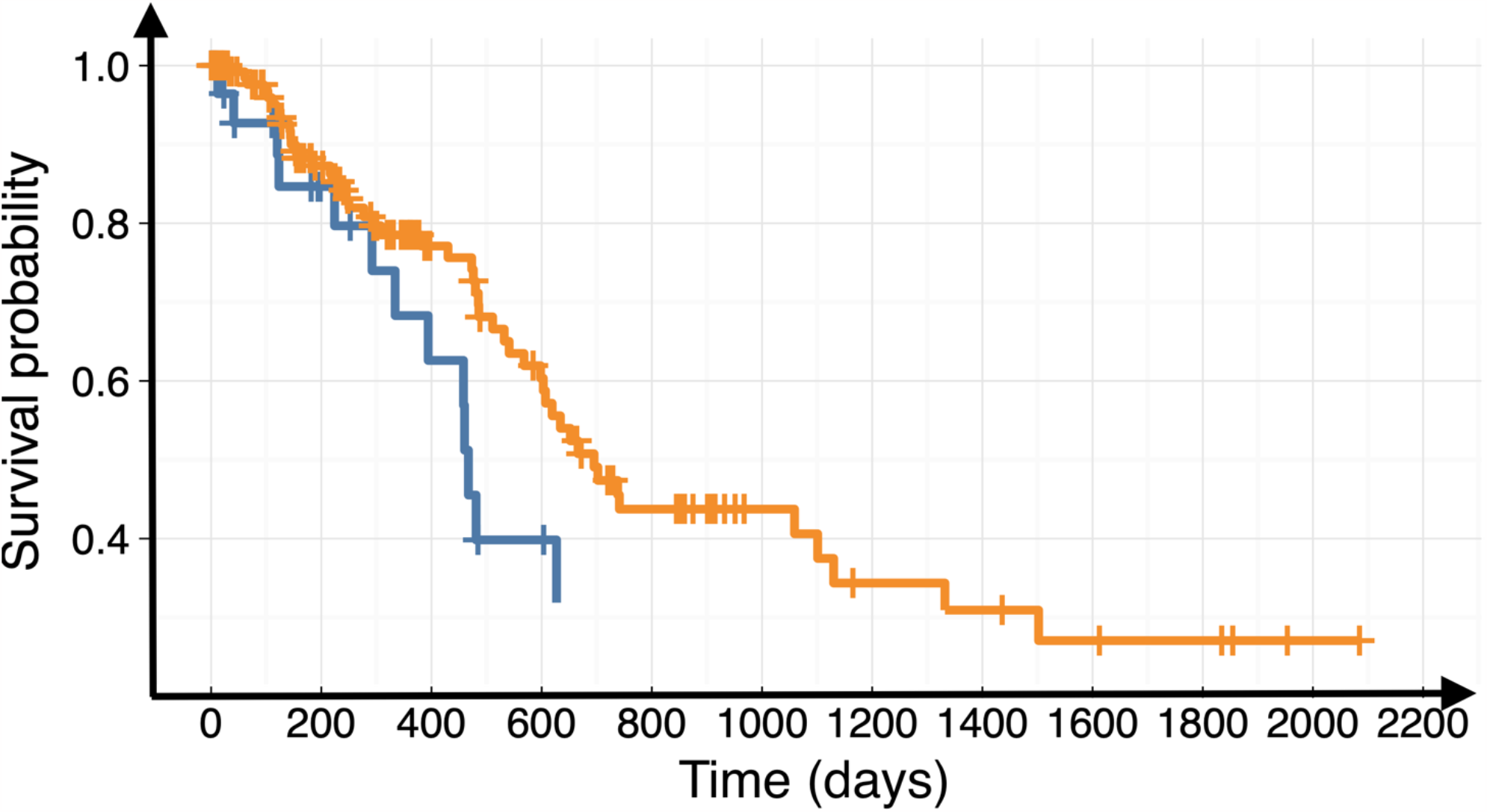
Survival curves for patients with different SNVs. Patients (blue) with somatic mutation on genes, *CDKN2A, PRKG1*, and *BCORL1*, have shorter overall survival time than other pancreatic cancer patients (orange).

In this study, we explored a genome-wide and radiome-wide association investigation. Screening hundreds of thousands of genetic variants across the entire genome, genome-wide association studies (GWAS) have been widely used to identify disease-specific genetic variants and use them in broad clinical and biological applications. For pancreatic cancer, previous GWAS have identified new and useful risk loci^47^. For this extremely lethal disease, these types of findings can be especially important owing to the complexity as well as the dynamic heterogeneity over the rapid progression of the disease. However, GWAS are expensive to conduct and require tissue samples that are not readily available. For pancreatic cancer, this is especially the case because tissue samples are usually only procured during surgery owing to the high risks associated with biopsy, and only around one-fifth of all pancreatic cancer patients are operable due to late stages at detection^48^. The association study approach described in this work can help link the radiome with the genome, i.e., link phenotypic radiomics information from medical images with genotypic information, therefore enabling a much broader and longitudinal genome-wide search for this highly dynamic disease.

The Pancreatic Cancer Rapid Autopsy provides a unique dataset that allows comprehensive investigation of pancreatic cancer with a systems approach. In this proof-of-concept study, a genome-wide and radiome-wide association analysis was conducted on whole-exome sequencing data from both primary tumor and normal pancreatic tissue from the rapid autopsy. Our approach normalizes the SNVs identified on tumor tissue by those on normal tissue, i.e. the approach teases out all somatic SNVs and selects only tumor SNVs. This way each patient acts as their own control, thereby suppressing the immense background noise and focusing only on the tumor-specific genomic signals. Potentially, similar “normalization” approaches could be applied to zoom in on the genomic changes between the primary tumor and each of the metastatic tumors, and the temporal changes of all tumors over the course of disease progression and treatments. For the former, the large tissue collection from the rapid autopsy is uniquely valuable by providing primary and all metastatic lesions as well as normal organ tissues. This radiome-genome association approach we established in this work could facilitate these investigations with relevant radiomics features. For the latter, direct genomic assessment is not possible as tissue samples cannot be collected repeatedly along the time course. On the other hand, because periodical medical imaging is already part of the cancer care routine, the relevant radiomics features can be used as surrogates to assess the longitudinal genetic changes accompanying the rapid and heterogeneous progression of this vicious disease. Together, this approach and the future investigations it enables may help decipher mechanisms and pathways of how pancreatic cancer cells progress and respond to treatments and shed light on better treatment options.

Radiogenomics is an existing branch of radiomics^49^. By combining genomic data and imaging features, it has been shown to yield imaging biomarkers and provide valuable information for diseases, especially cancer. However, among different cancers, there is a relative paucity of radiogenomics literature for pancreatic cancer, largely due to the limited known molecular markers for this highly heterogeneous disease and the difficulty to obtain simultaneous imaging data and genomic data in well-synchronized time points for this rapidly progressing disease. Furthermore, all existing radiogenomics studies, including those on the pancreatic cancer^50^, focused only on known molecular markers. In contrast, our genome-wide and radiome-wide association study applies a system-wide search through large-scale radiomics and genomics data to explore novel imaging and genomic biomarkers and mechanisms. In this proof-of-concept preliminary study, radiomics and primary tumor whole genome information were correlated for pancreatic cancer patients. Using the last pancreatic CT scan of the patient, the imaging date was reasonably close to the date of death when the tissues used for genomic investigations were collected through rapid autopsy comparable in quality to that obtained by surgical resection.

While the concept of a system-wide large-scale radiome- and genome-wide association study is innovative and the results encouraging, the study is not without limitations. First, the cohort size is rather small with only 26 patients. The size is limited by data availability and more so by the substantial cost associated with the whole-exome sequencing of each tumor and tissue sample. This cost highlights the potential benefits of this multiomcis association approach to develop imaging surrogates for these costly large-scale screenings. At the same time, as our study applied the somatic SNVs of each patient as their own control, tumor SNVs can be identified with much higher specificity in even a small cohort. Another limitation is the slight heterogeneity of the imaging data in terms of CT scanner model and acquisition protocol, as well as the time point of the scan relative to tissue sample collection. These variances are inevitable as the unique dataset came from the Pancreatic Rapid Autopsy Program which is retrospective and curated carefully over more than a decade of time. On the other hand, coming from a single institution, the CT scanners used for this study were all from a single vendor and on a single line of models, and the imaging protocols were largely similar. The imaging dates for the cohort ranged from 1 to 224 days before patient death but were relatively synchronized with the tissue collection with imaging dates within 1 or 2 months for most patients. For large-scale omics research, false positives are always a potential challenge, especially when the sample size is small. In our study, we employed some strategies to minimize false positives, such as applying the gene-based burden-testing approach and selecting a stricter significance level at P-value = 10^−4^.

Future applications of this novel methodology could include genome and radiome association studies on metastatic lesions to investigate their similarities and differences with the primary pancreatic tumor in terms of these molecular and imaging metrics. This could further validate the new approach and understand the complex tumor mutations that occur during the progression of pancreatic cancer. The application of the discovered imaging biomarkers to additional patients as well as the longitudinal images of these patients would be an additional investigation of value. The latter may shed light on the temporal changes of relevant genomic markers as the disease progresses and the patient responds to treatment.

## Methods and Materials

### Study population

Patients were included in this study from a unique database of the University of Nebraska Medical Center Pancreatic Cancer Rapid Autopsy Program. For over a decade, the program has been collecting large quantities of tumor and tissue samples from autopsies performed within hours of patient demise. In the rapid autopsy, all primary and metastatic tumors, and a large number of tissue samples such as liver, lung, spleen, and kidney are collected under rapid conditions that produce tissue that is comparable in quality to that obtained by surgical resection. The resected autopsy samples are reviewed and annotated by at least two pathologists, in concert with lab members who conduct the autopsies. Twenty-six patients from the pancreatic cancer rapid autopsy program with comprehensive, unique tissue sample collection and longitudinal contrast-enhanced CT images were included in this study. All data collection was approved by the IRB of our institution (Protocols: 728-16-EP and 127-18-EP).

### DNA isolation and whole exome sequencing (WES)

DNAs were extracted from tumor tissues and from healthy tissues in the liver, kidney etc. Illumina TruSeq DNA Exome kit was used for exon capture. Sequencing was carried out using Illumina 2×100 bp paired-end sequencing on a HiSeq 2500 instrument according to the manufacturer’s recommendation.

### Genome-wide identification of somatic single-nucleotide variants (SNVs)

When applied to many samples of the same cancer type, the identification of the cancer driver gene can be conducted to search for multiple recurrences of somatic mutations in the same gene. With the WES data for the tumor and tumor-free organ tissues from 26 patients, tumor-specific somatic SNVs were identified with VarScan2 ^51^ after the standard read preprocessing and read-mapping by BWA^52^. Based on the FDR-adjusted P-values calculated by VarScan2, we retained the significant somatic mutations with a cut-off of the adjusted P-value < 10^−5^ for subsequent analyses.

### Imaging Studies

For the 26 patients included in the study, varying numbers (3-30) of contrast-enhanced abdominal CT scans were acquired per standard pancreatic cancer care from diagnosis to longitudinal monitoring, using Lightspeed VCT, Lightspeed Pro 16, or Lightspeed RT16 (GE Healthcare, Boston, Massachusetts, USA). For the image acquisition, patients received ISOVUE injection with bolus triggering arterial phase imaging about 30 s and venous phase about 60 s after injection. These scans used slice thickness of 1.5-5 mm, with an in-plane resolution of 0.6-0.8 mm. For the purpose of this study, the last available CT scan prior to the patient’s death was used for radiomics analysis for the patient. This way we could get the closest match between radiomics information from the imaging and genomic information from the rapid autopsy.

### Radiomics Feature Extraction and Selection

Pancreatic tumor volume-of-interest (VOI) was manually segmented by two experienced clinical investigators using a consistent window/level setting and reconciled disagreements to mitigate intra- and inter-observer uncertainty. From each tumor, VOI, 944 radiomics features were extracted using the radiomics module on 3D Slicer (version 4.10)^53^ and visualized using an interactive visualization platform. A resampled 2×2×2 mm^3^ voxel size and a bin width of 25 were used for feature extraction. The features are defined in compliance with feature definitions as described by the Imaging Biomarker Standardization Initiative (IBSI)^54^ and can be divided into original features (107 features), Laplacian of Gaussian features (LoG, 93 features) and wavelet features (744 features). The original features can be subdivided into 6 classes, including 14 Shape features, 18 First Order statistical features, 38 Gray Level Dependence Matrix (GLDM) features, 16 Gray Level Run Length Matrix (GLRLM) features, 16 Gray Level Size Zone Matrix (GLSZM) features, and 5 Neighboring Gray Tone Difference Matrix (NGTDM) features. The wavelet features included all except Shape features calculated on the filtered images with all 8 combinations of applying either a High or a Low pass filter in each of the three dimensions. All features are pre-selected to eliminate features unstable to respiratory motion and inter-observer contouring uncertainty^55^. The stable features were subsequently selected again using a recursive correlation pruning step to remove redundant features. We focused on 170 nonredundant radiomics features, which include tumor diameters, tumor major axis length, tumor flatness, tumor elongation, etc.

### Genome-wide association analysis between radiomics features and somatic mutations

Based on our discovered genes with high reoccurrences rate of somatic mutations and radiomics features from the corresponding tumor, we conduct an association study between these genes and radiomics features of CT scans from the same population. Here, we employ the gene-based burden-testing approach. For this approach, we use the number of individuals carrying variants in each gene to associate with traits in cohorts^56^. We applied the sequence kernel association test (SKAT) ^57^ to test the association between each radiomics feature and somatic mutations within each gene. SKAT was originally designed for testing the association between a trait and the rare variants in a genomic region and was based on a variance-component score test in a mixed-model framework. It is shown to have much higher power than many other burden tests for gene-based GWAS. We focus on the 132 genes with somatic mutations in at least three patients (the specific mutations can be different among these patients) and tested their association with each of the 944 radiomics traits. The top two PCs of the somatic mutation matrix are used as covariates in the null model. In our dataset, the reoccurrences of individual somatic mutations are generally low due to the low sample size. Aggregating their potentially heterogeneous effects using SKAT is expected to improve the detection power. The output p-values are adjusted for multiple testing based on Benjamini-Hochberg Procedure.

### TCGA data and survival analysis

The R package, RTCGA, (https://rtcga.github.io/RTCGA/) and the full set of somatic mutations discovered by TCGA data for the cohort of Pancreatic Cancer from Firehose (https://gdac.broadinstitute.org/)^46^ were used to get mutation and survival information of 185 Pancreatic Cancer patients for survival analysis. The function, *kmTCGA*(), in RTCGA was used to plot Kaplan-Meier estimates of survival curves for survival data from patients with or without given mutations. The function, *pairwise_survdiff()*, in the R package of survminer was used to have the comparisons of multiple survival curves.

## Data Availability

All data produced in the present work are contained in the manuscript.

## Data availability

The data of all somatic SNVs and radiomics features that were generated and analyzed during this study are included in this published article and the Supplementary Information files.

## Author contributions

D.Z., C.Z., and M.H. conceived the project. M.B. and A.K. performed expert contouring. P.G., D.Z., Y.S., Q.D., J.W., S.B., and K.P. curated the data. C.Z., D.Z., Q.Z., X.L., H.Y., and H.D. performed data analysis. C.Z., D.Z., and M.H. drafted the manuscript. M.H. provided expert knowledge and funding support. All authors reviewed and approved the manuscript.

## Supplementary Tables

**Supplementary Table S1: Somatic SNVs obtained from WES**.

**Supplementary Table S2: Recurrence rates of SNVs in the top recurrent genes**.

**Supplementary Table S3: Radiomics features of tumor CT images for patients**.

